# Development and validation of a population pharmacokinetic model to guide perioperative tacrolimus dosing after lung transplantation

**DOI:** 10.1101/2023.06.26.23291248

**Authors:** Todd A. Miano, Rui Feng, Stephen Griffiths, Laurel Kalman, Michelle Oyster, Edward Cantu, Wei Yang, Joshua M. Diamond, Jason D. Christie, Marc H. Scheetz, Michael G. S. Shashaty

## Abstract

**Background:** Tacrolimus therapy is standard of care for immunosuppression after lung transplantation. However, tacrolimus exposure variability during the early postoperative period may contribute to poor outcomes in this population. Few studies have examined tacrolimus pharmacokinetics (PK) during this high-risk time period.

**Methods:** We conducted a retrospective pharmacokinetic study in lung transplant recipients at the University of Pennsylvania who were enrolled in the Lung Transplant Outcomes Group (LTOG) cohort. We derived a model in 270 patients using NONMEM (version 7.5.1) and examined validity in a separate cohort of 114 patients. Covariates were examined with univariate analysis and multivariable analysis was developed using forward and backward stepwise selection. Performance of the final model in the validation cohort was examined with calculation of mean prediction error (PE).

**Results:** We developed a one-compartment base model with a fixed rate absorption constant. Significant covariates in multivariable analysis were postoperative day, hematocrit, transplant type, *CYP3A5* genotype, total body weight, and time-varying postoperative day, hematocrit, and CYP inhibitor drugs. The strongest predictor of tacrolimus clearance was postoperative day, with median predicted clearance increasing more than threefold over the 14 day study period. In the validation cohort, the final model showed a mean PE of 36.4% (95%CI 30.8%-41.9%) and a median PE of 7.2% (IQR −29.3%−70.53%).

**Conclusion:** Postoperative day was the strongest predictor of tacrolimus exposure in the early post-lung transplant period. Future multicenter studies employing intensive sampling to examine a broad set of variables related to critical illness physiology are needed to understand determinants of clearance, volume of distribution and absorption in this population.

## Introduction

Tacrolimus is a potent calcineurin inhibitor that is standard of care for immunosuppression after lung transplantation (1,2) Despite routine use of daily therapeutic drug monitoring (TDM) in the early postoperative period, tacrolimus exposure during this vulnerable time frame is highly variable and may contribute to poor outcomes in this population (3,4). Tacrolimus is characterized by high inter- and intra-patient variability of pharmacokinetic (PK) parameters and a narrow therapeutic index. High concentrations can cause renal vasoconstriction (5) that may lead to acute kidney injury (AKI), which occurs in 50-70% of patients postoperatively, and is linked to increased early mortality as well as higher chronic kidney disease (CKD) risk in survivors (4,6,7). Low concentrations during the postoperative period may increase the risk for acute cellular rejection (ACR), which is especially concerning given that up to 30-50% of lung transplant recipients may experience ACR during the first year after transplant (8,9). Interventions to optimize early tacrolimus exposure thus constitute a significant unmet need in the lung transplant population.

Rather than the fixed initial dose approach employed by most centers (2), a proactive strategy would leverage information on tacrolimus pharmacokinetics to guide personalized dose initiation and titration. However, such approaches are hindered by limited data on the determinants of tacrolimus pharmacokinetics in lung transplant recipients, particularly during the early postoperative period. In the kidney transplant population, the dominant impact of allelic variation in the expression of the cytochrome P450 3A5 (CYP3A5) drug metabolizing enzyme has been studied extensively, with genetic variants causing higher CYP3A5 gene expression linked to approximately 50% higher tacrolimus dosing requirements (10). However, we have shown that CYP3A5 genotype explains only 19% of the variability in tacrolimus concentration-dose ratio (CDR) during the early postoperative period after lung transplantation, a time when clinical factors may have a much larger impact (4). Compared to kidney transplantation, lung transplantation is a much greater physiologic insult in which all patients are critically ill postoperatively, requiring mechanical ventilation and substantial hemodynamic support during the first few days after transplant. Patients frequently experience gut dysmotility, altered splanchnic and hepatic blood flow, inflammatory mediated reductions of hepatic CYP enzyme activity, and rapidly fluctuating hemoglobin concentrations (3,11). These physiologic responses may have profound impacts on tacrolimus pharmacokinetics, representing substantial sources of residual pharmacokinetic variability that are rarely observed after kidney transplantation (11). Consequently, the generalizability of pharmacokinetic data between kidney transplant patients and lung transplant patients is low, particularly during the early postoperative period.

Although the pharmacokinetics of tacrolimus have been extensively investigated in kidney and liver transplant populations, few studies have developed population pharmacokinetic models in lung transplant recipients (3,12,13), with limited data on the early postoperative period (3). We thus aimed to develop and validate a tacrolimus population pharmacokinetic model that includes clinical and genetic determinants of tacrolimus response during the initial 14 days after lung transplantation.

## Methods

### Study design and population

We conducted a retrospective pharmacokinetic study in lung transplant recipients at the University of Pennsylvania who were enrolled in the Lung Transplant Outcomes Group (LTOG) study, a multicenter prospective cohort study of primary graft dysfunction (PGD) (14). LTOG data were merged with additional data obtained from our electronic health record (medications, tacrolimus concentrations, other laboratory values) for the current analysis. We included LTOG subjects who underwent single or bilateral lung transplantation between November 2008 and August 2018. Exclusion criteria were age less than 18 years, combined organ transplantation other than heart-lung (because of the limited number of these procedures), exposure to cyclosporine (the key alternative calcineurin agent to tacrolimus), exposure to intravenous tacrolimus, or missing tacrolimus concentration data. We randomly divided eligible patients into derivation and validation sets targeting an approximate 70%:30% split, respectively. Tacrolimus dosing, concentration, and covariate data were collected from the initial 14 days after transplantation. The study was approved by the University of Pennsylvania institutional review board and subjects provided informed consent for enrollment in the LTOG.

### Immunosuppression regimen

Induction immunosuppression consisted of high dose corticosteroids and basiliximab. Maintenance immunosuppression was initiated within 12-24 hours post-operatively, and consisted of tacrolimus, an antiproliferative agent (mycophenolate or azathioprine), and low dose corticosteroids (tapered to a target dose of 10-20 mg prednisone daily). Tacrolimus was administered twice daily orally or sublingually. Dosing was at the discretion of the treating clinicians and was not guided by any formal dosing algorithm. The target tacrolimus concentration during the first three months after transplant was 8-12 ng/ml and remained uniform during the study period. Daily trough concentrations were obtained prior to the morning dose during the first two post-operative weeks, with less frequent monitoring employed as maintenance doses were stabilized. Tacrolimus whole blood concentrations were measured using a standard liquid chromatography-mass spectrometry technique (15).

### Population pharmacokinetic analysis

We performed population PK analysis using NONMEM (version 7.5.1) through the interface provided by PDx-POP (version 5.3, ICON plc, Leopardstown, Dublin, Ireland). Output was summarized using R (version 3.5.1, R Foundation for Statistical Computing, Vienna, Austria, www.r-project.org) and Stata (version 17.0, StataCorp, College Station, Texas, USA). All models were run with the First-Order Conditional Estimation with Interaction method.

### Base model development

We *a priori* specified a one-compartment structural model for this sparse dataset consisting mainly of trough concentrations, as the lack of concentration data in the early time period after tacrolimus administration precluded estimation of intercompartmental transfer (16). Similarly, the absorption rate constant (ka) was fixed to 4.5/h (16), as it cannot be reliably estimated from trough data alone. Because bioavailability (F) could not be estimated, parameter values were estimated as ratios (CL/F, Vd/F). We described random interindividual variability using an exponential variance model, with individual random effects for all parameters assumed to follow a multivariate normal covariance structure. Residual variability was evaluated using an additive and proportional error model.

### Patient-level covariate-based model development

After base model development, we examined a series of candidate covariates that were chosen for screening *a priori* based on findings from our previous study (4), other tacrolimus pharmacokinetic literature (3,11,16), and physiologic plausibility. Genotyping was performed on peripheral blood samples using an Affymetrix Axiom genotyping array with Applied Biosystems Axiom 2.0 Reagents (17). We focused on single nucleotide polymorphisms (SNPs) that have been consistently linked to tacrolimus dose response in previous studies. CYP3A5 is the predominant tacrolimus metabolic pathway in patients that express the enzyme and previous research has shown this enzyme to be the most important source of genetic variation in tacrolimus dose response (10). We examined CYP3A5 variants rs776746 [*CYP3A5*3*, (6986A>G in intron 3)]; rs10264272 [*CYP3A5*6*, (14690G>A in exon 7)]; and rs41303343 [*CYP3A5*7*, (27131-27132insT)]. Prior research has shown that the *6 and *7 variants produce similar loss of CYP3A5 activity compared to the *3 variant, but are more commonly found in the African American population (10). We classified CYPA5 activity by drawing on all three SNPs as recommended by the Clinical Pharmacogenetics Implementation Consortium (CPIC) (10): extensive metabolizers (CYP3A5*1*1), intermediate metabolizers (CYP3A5*1*3, *1*6, *1*7), poor metabolizers (CYP3A5*3*3, *6*6, *7*7, *3*6, *3*7, *6*7). Given a small number of patients with the extensive metabolizer genotype, we combined extensive and intermediate metabolizers into one group. In addition, we genotyped CYP3A4*22 (rs35599367, g.15389C>T in intron 6), based on prior data showing that this SNP has important effects on tacrolimus dose response after controlling for the effects of CYP3A5 (18,19).

We included clinical variables such as demographics and pre-transplant health status (age, weight, race, sex, cystic fibrosis diagnosis); potential drug-drug interactions between tacrolimus and CYP enzyme inhibitors commonly administered in lung transplant patients (fluconazole, voriconazole, amiodarone), modeled as time-varying covariates lagged by 24 hours; hematocrit, modeled as a time-varying covariate, which is a marker for tacrolimus binding to red blood cells (11,20,21); and markers of post-operative severity of illness (transplant type [single vs. bilateral], post-operative day, and the occurrence of primary graft dysfunction (PGD) (22) during the first three post-operative days). Given the dynamic nature of patient physiology during the postoperative period, we anticipated the effect of transplant type and PGD to vary over time as patients recover from the surgical insult. We thus modeled these variables as both time-invariant effects, and as time-varying effects (i.e., estimating separate coefficients on clearnace during postoperative days 0-3 and 4-14). Although post-operative corticosteroid dosing may induce tacrolimus metabolism (23), we did not examine this variable due to a lack of variability in exposure (all patients received corticosteroids); and because CYP metabolism induction typically has a delayed onset of 7-14 days (23). Age and cystic fibrosis were strongly correlated, as were race and CYP3A5 genotype. We selected cystic fibrosis and CYP3A5 for model inclusion, as these variables have the most apparent biologic relationship with tacrolimus metabolism (13).

We first examined covariate relationships with CL and VD graphically. We then added covariates to the base model in a univariate fashion to identify those significantly improving model fit (decrease in objective function value (OFV) of at least 3.84, which corresponds to p-value <0.05). We then conducted a manual forward stepwise multivariable analysis with significant covariates from univariate screening. In this step, we retained covariates in the model that decreased OFV by at least 3.84. We then manually applied a backward elimination procedure to the full model, retaining covariates that resulted in an increase of OFV of at least 6.64 (p-value<0.01) upon removal.

We coded discrete covariates as 1 indicating presence of the covariate and 0 indicating absence of the covariate and applied a multiplicative function specified as TVP = θ_TVP_ × (θ_cov_)^COV^, where TVP is the typical value of the parameter, θ_TVP_ is the population parameter estimate and θ_cov_ is the effect of the covariate (24). Thus, in the absence of the covariate, (θ_cov_)^COV^ is equal to 1 and there is no effect on the parameter. We evaluated continuous covariates with normalized power models, specified as TVP = θ_TVP_ × (covariate value/covariate)^θcovariate^, where the covariate value is the value at the time the pharmacokinetic samples were obtained, covariate_med_ is the median value of the covariate in the population, and θ_covariate_ is the effect of that covariate on the parameter of interest (24).

### Model evaluation in the derivation cohort

We assessed model adequacy using goodness-of-fit criteria including visual inspection of diagnostic scatter plots (observed vs predicted concentration, observed and predicted concentration vs time, and weighted residual vs predicted concentration or time), plausibility of parameter estimates based on prior literature (16), precision of parameter estimates as measured by asymptotic standard errors, successful model convergence, changes in Akaike Information Criteria (AIC), and the size of interindividual and residual variabilities for the specified model. Lastly, we calculated shrinkage for model parameters with interindividual variability estimates, considering values below 25% acceptable (25).

### Model evaluation and dosing simulations in the validation cohort

We used coefficients from the final derived model to calculate predicted concentrations using the tacrolimus dosing and covariate data in the validation cohort. Predicted concentrations were then compared with observed concentrations in the validation cohort. We calculated prediction error (PE) as a measure of predictive performance (26), defined as PE = (C_pred_ − C_obs_)/C_obs_ × 100%, where C_pred_ represents the population predicted concentration and C_obs_ represents the observed concentration in the validation cohort. We summarized PE with mean (95% CI) and median (interquartile range). We *a priori* specified successful validation of the final model as showing both mean and median PE values to be less than 20%. Additionally, we calculated the percentage of predicted concentrations that were within 2 ng/mL and 4 ng/mL of observed concentration as a measure of clinical utility, as the range of early post-lung transplant target concentrations targeted by clincians varies widely (2). Lastly, model performance was visualized with a plot of the population predicted concentrations versus observed concentrations.

## Results

### Cohort characteristics

During the study period, 384 patients (derivation n=270, validation n=114) were enrolled in the LTOG study, genotyped, and had at least one tacrolimus concentration obtained postoperatively. Patient characteristics are shown in Table 1. The average age was 60 years and approximately two-thirds of patients in each cohort underwent bilateral lung transplantation. One in five patients had a CYP3A5 intermediate or extensive metabolizer genotype, and exposure to potentially interacting drugs was common. Median inpatient follow-up was 13 days in both cohorts and the median number of tacrolimus trough concentrations per patient was 13 in the derivation cohort and 12 in the validation cohort (approximately one concentration per day). The median initial tacrolimus daily dose was 2 mg in both cohorts. The individual trend lines of tacrolimus concentrations in both cohorts are shown in Figure 1, showing substantial variability relative to the target concentration range.

**Figure 1.**
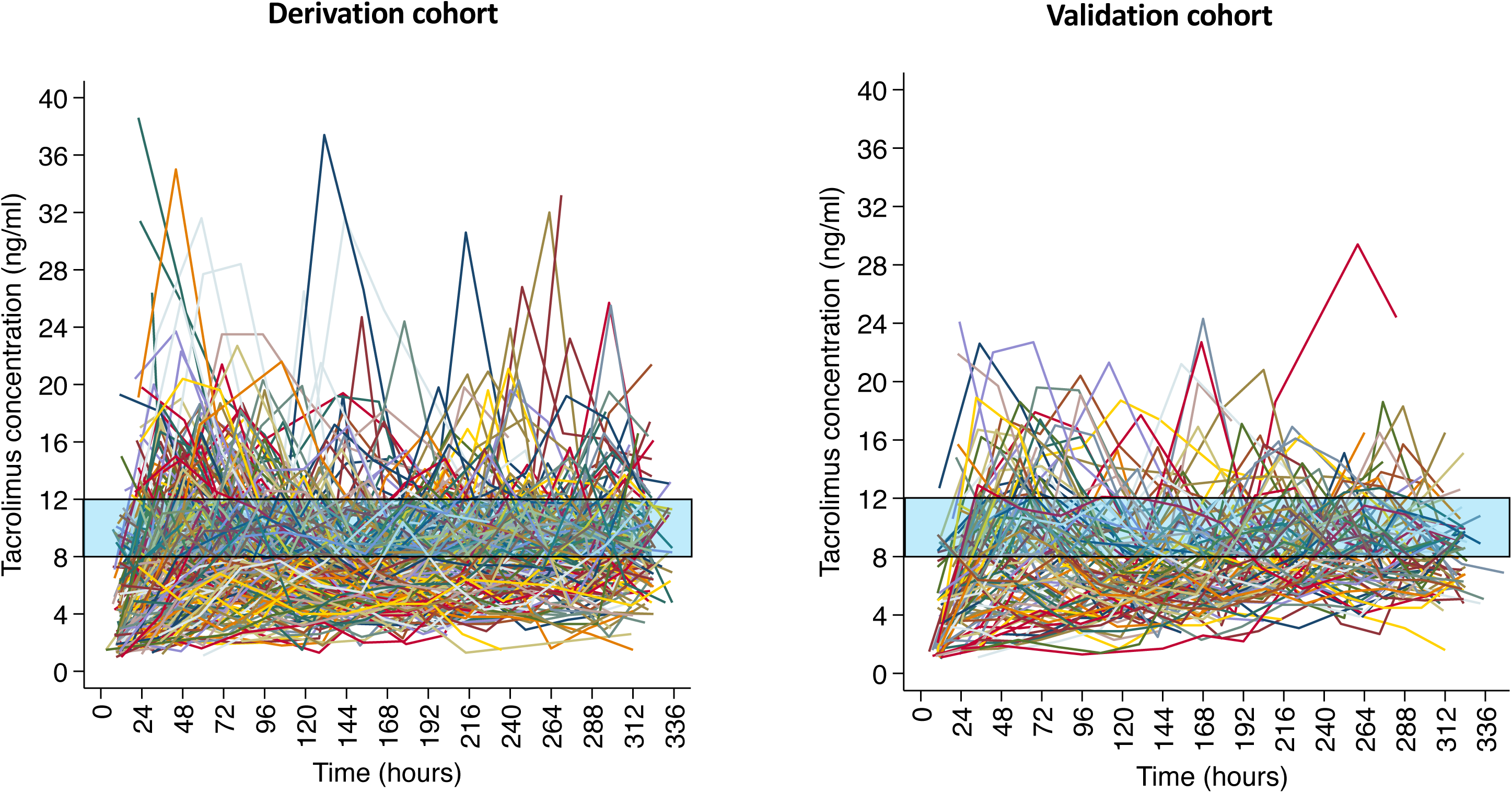
Individual trend lines of tacrolimus concentration over time in the derivation and validation cohorts. The shaded area represents the target tacrolimus concentration range (8–12 ng/mL).

**Table 1.**

|  | **Derivation cohort**<br>(n=270) | **Validation cohort**<br>(n=114) |
| --- | --- | --- |
| Age, years, median (IQR) | 61 (51, 66) | 60 (52, 65) |
| Female sex, n (%) | 107 (40) | 44 (39) |
| Weight, kg, median (IQR) | 74 (59, 88) | 77 (61, 87) |
| Race, n (%) |  |  |
| White | 237 (88) | 98 (86) |
| African American | 23 (9) | 10 (9) |
| Other | 10 (3) | 6 (5) |
| Cystic fibrosis, n (%) | 29 (11) | 8 (7) |
| Transplant type, bilateral, n (%) | 177 (66) | 78 (68) |
| Primary graft dysfunction, n (%)<sup>a</sup> | 61 (23) | 33 (30) |
| Hematocrit, (%), median (IQR) | 33 (30, 38) | 34 (30, 38) |
| CYP3A5 metabolizer genotype, n (%)<sup>b</sup> | 53 (20) | 22 (19) |
| CYP3A4 metabolizer genotype, n (%)<sup>c</sup> | 29 (7) | 10 (9) |
| CYP enzyme inhibitors, n (%)<sup>d</sup> |  |  |
| Voriconazole | 95 (35) | 33 (29) |
| Fluconazole | 20 (7) | 4 (4) |
| Amiodarone | 96 (36) | 43 (38) |
| Follow-up duration, days, median (IQR) | 13 (11, 14) | 13 (10, 14) |
| Tacrolimus concentrations |  |  |
| Total samples | 3143 | 1279 |
| Samples per patient, count, median (IQR) | 13 (10, 14) | 12 (10, 13) |
| Initial tacrolimus dose, mg, median (IQR) | 2 (2, 2) | 2 (2, 2) |

a- diagnosed by postoperative day three or earlier; b- presence of at least one allele encoding an extensive or intermediate metabolizer phenotype as defined by the Clinical Pharmacogenetics Implementation Consortium consensus guidelines (10); c- presence of at least one allele encoding a poor metabolizer phenotype; d- ever exposed during the fourteen day study period; IQR – interquartile range

### Model building steps

#### Base model

Parameters of the one-compartment base model with a fixed rate absorption constant are shown in Table 2. Goodness of fit plots are shown in Figure 2, with plots of predicted versus observed stratified by time (postoperative days 0-3 and 4-14). The base model tended to under predict observed concentrations during the early time period, while overpredicting observed concentrations during the later time period.

**Figure 2.**
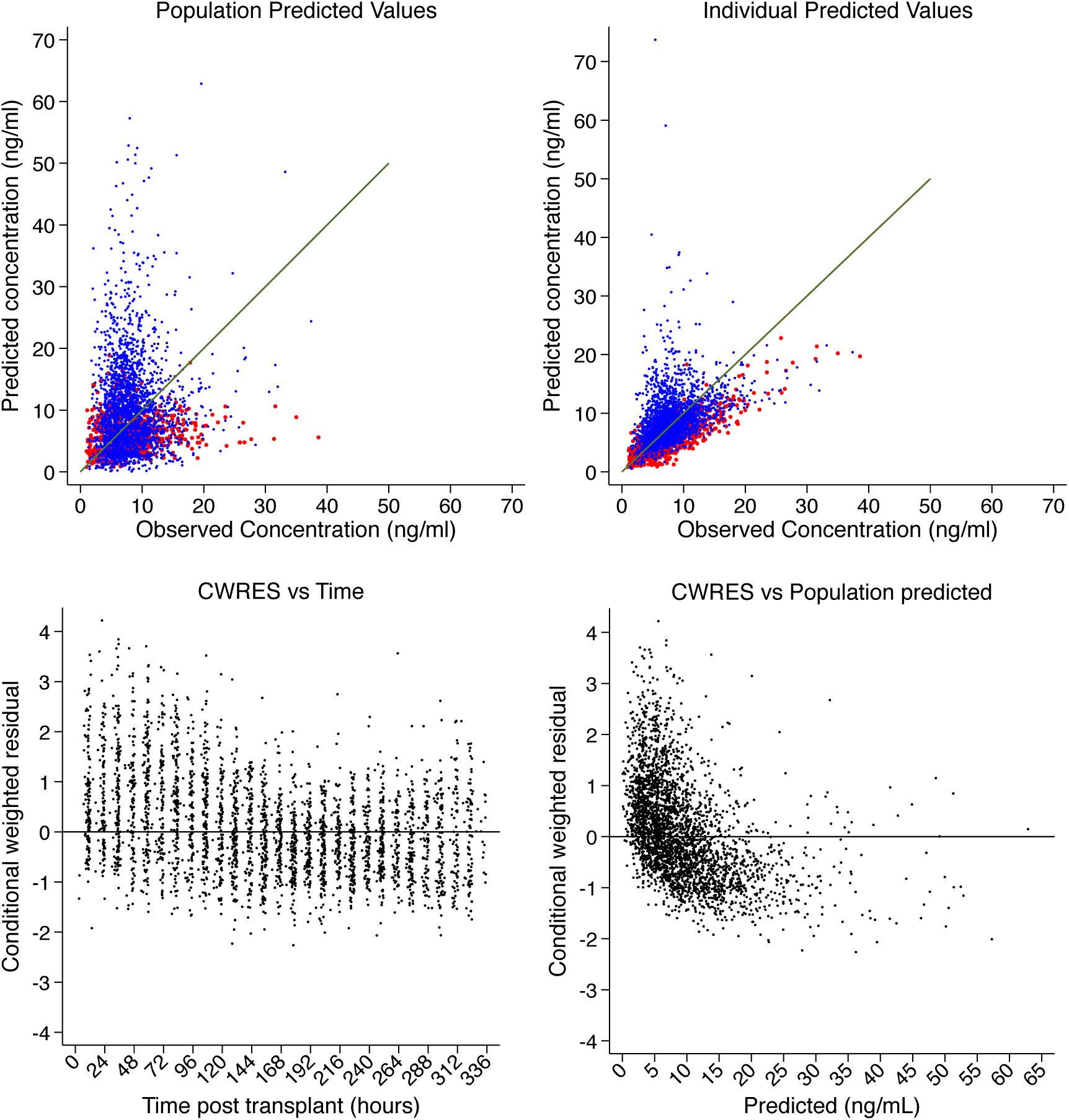
Diagnostic plots of the base model in the derivation cohort. Observed vs. predicted plots stratified by postoperative day (red days 0-3, blue days 4-14). CWRES-conditional weighted residual error.

**Table 2.**
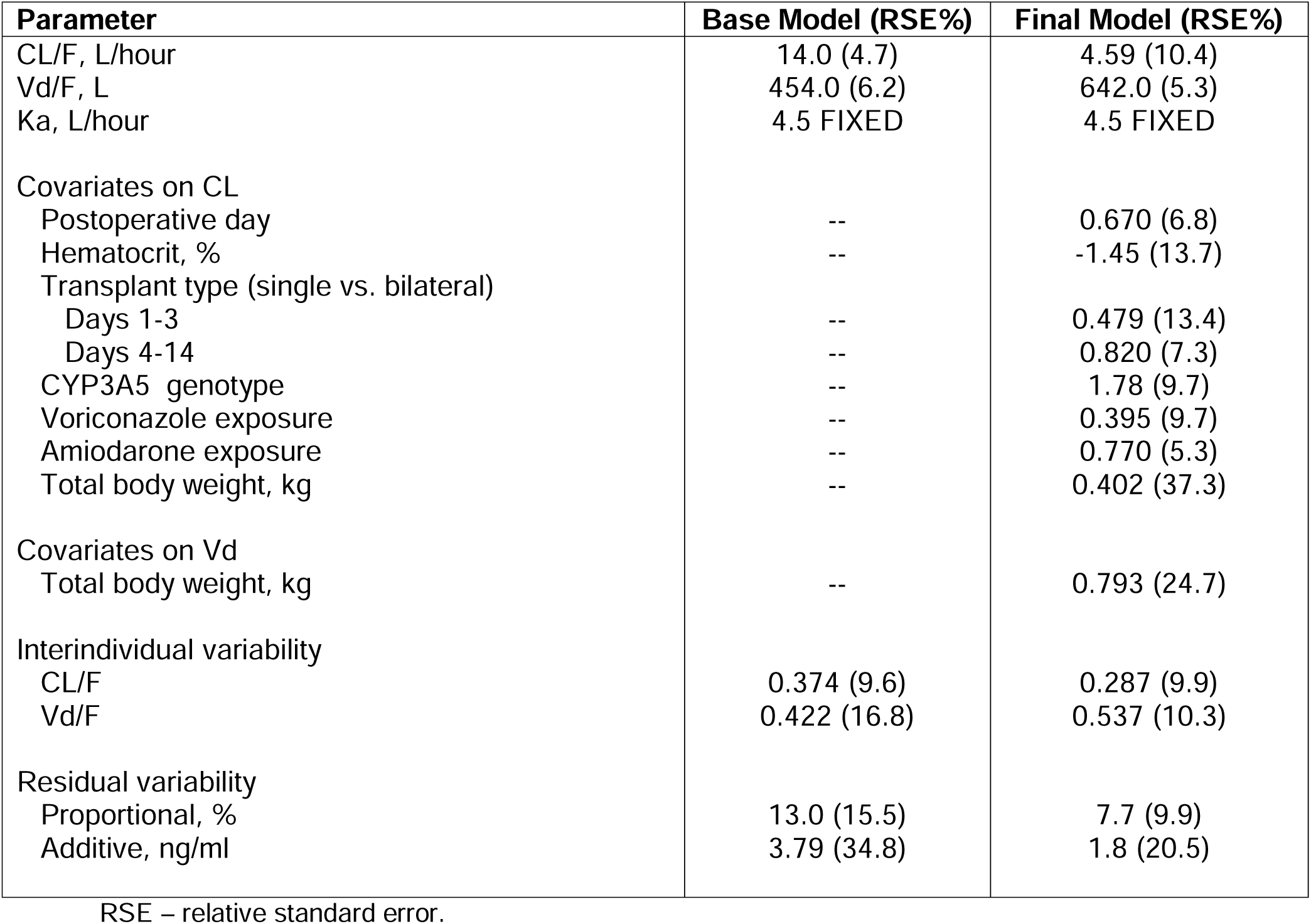
Parameter estimates of the base model and final model

#### Model incorporating patient-level covariates

Results of patient-level covariate modeling steps are shown in Table 3. Significant covariate effects on CL in univariate analysis included total body weight, CYP3A5 and CYP3A4 genotypes, transplant type, postoperative day, PGD, hematocrit concentration, and concomitant use of CYP inhibitors (voriconazole, fluconazole, and amiodarone). Total body weight was also significantly associated with Vd. Transplant type and PGD significantly improved model fit when specified as time varying effects (i.e., separate coefficients for days 0-3 and days 4-14), but did not significantly improve model fit as time invariant effects (i.e., a single coefficient for days 0-14). After forward and backward stepwise selection, the final model included postoperative day, hematocrit, transplant type, CY3A5 genotype, enzyme inhibitor drugs, and total body weight on Cl and Vd (Table 2). The strongest predictor of tacrolimus clearance was postoperative day, with median predicted clearance increasing more than threefold over the 14 day study period (Figure 3).

**Figure 3.**
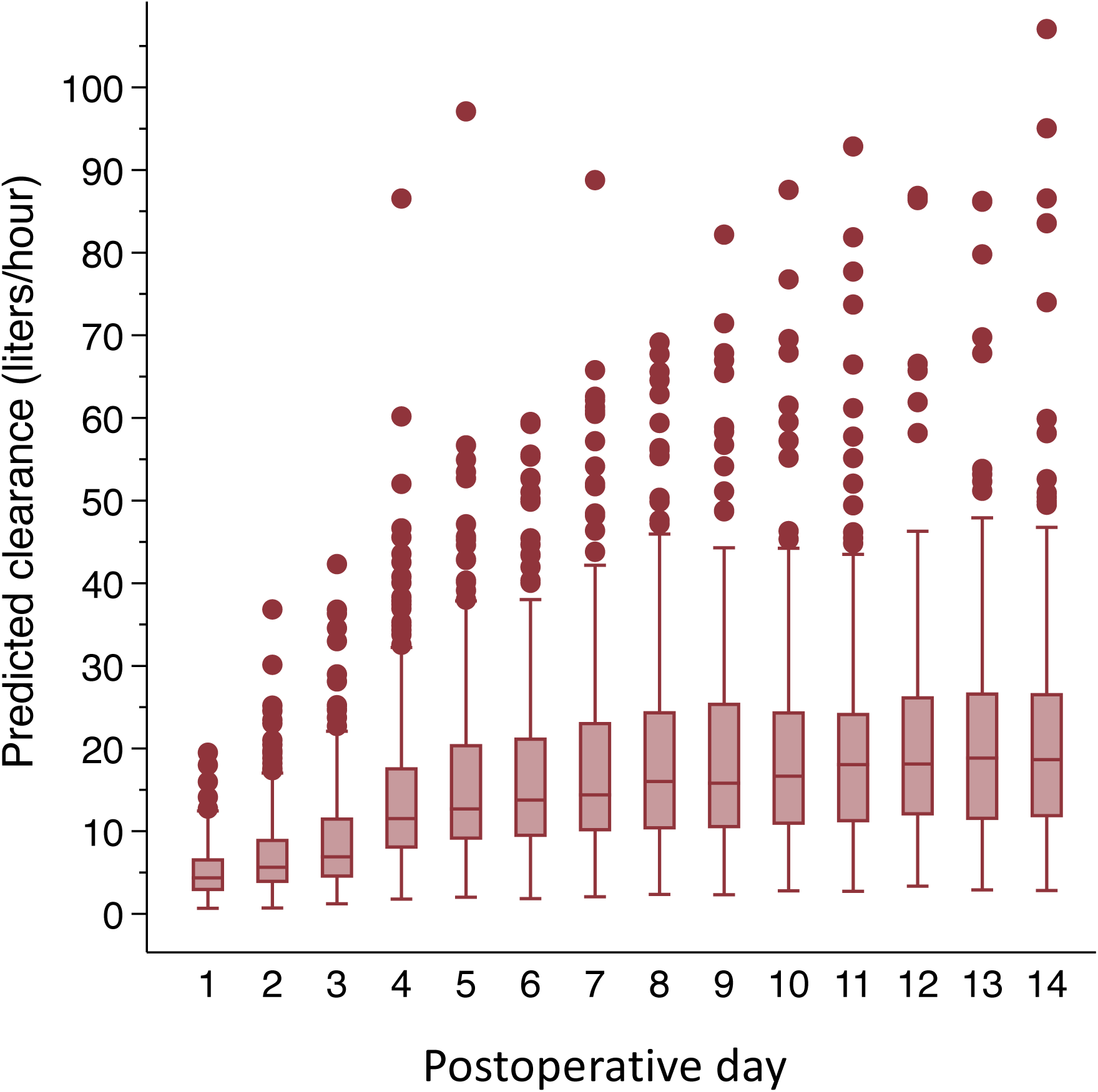
Diagnostic plots of the final model in the derivation cohort. Observed vs. predicted plots stratified by postoperative day (red days 0-3, blue days 4-14). CWRES-conditional weighted residual error.

**Table 3.** Model building steps

| **Covariate** | **OFV** | **Delta OFV** | **AIC** | **Delta AIC** |
| --- | --- | --- | --- | --- |
| Base Model | 12086.34 | -- | 12100.30 | -- |
| **Univariate covariate effects on clearance (CL)** |  |  |  |  |
| Total body weight | 12079.71 | -6.63 | 12095.70 | -4.60 |
| Postoperative day | 11107.33 | -979.01 | 11123.30 | -977.00 |
| Transplant type (single vs. bilateral) |  |  |  |  |
| Time invariant effect | 12083.47 | -2.87 | 12099.50 | -0.80 |
| Time varying effect (early vs. late) | 11509.63 | -576.71 | 11527.60 | -572.70 |
| PGD |  |  |  |  |
| Time invariant effect | 12086.26 | -0.08 | 12102.30 | 2.00 |
| Time varying effect (early vs. late) | 11945.08 | -141.26 | 11963.10 | -137.20 |
| CYP3A5 genotype | 12042.01 | -44.33 | 12058.00 | -42.30 |
| CYP3A4 genotype | 12078.81 | -7.54 | 12094.80 | -5.50 |
| Cystic fibrosis | 12086.24 | -0.10 | 12102.20 | 1.90 |
| Voriconazole coexposure | 12053.02 | -33.32 | 12069.00 | -31.30 |
| Fluconazole coexposure | 12076.32 | -10.02 | 12092.30 | -8.00 |
| Amiodarone coexposure | 12059.75 | -26.59 | 12075.70 | -24.60 |
| Hematocrit | 11849.44 | -236.90 | 11865.40 | -234.90 |
| **Univariate covariate effects on volume of distribution (Vd)** |  |  |  |  |
| Weight | 12063.97 | -22.37 | 12080.00 | -20.30 |
| **Multivariable models: forward selection** |  |  |  |  |
| Model 1: postoperative day on CL | 11107.36 | -- | 11123.40 | -- |
| Model 2: Model 1 + hematocrit on CL | 11025.08 | -82.28 | 11043.10 | -80.30 |
| Model 3: Model 2 + transplant type on CL | 10949.50 | -75.58 | 10971.50 | -71.60 |
| Model 4: Model 3 + CYP3A5 genotype on CL | 10916.60 | -32.90 | 10940.60 | 30.90 |
| Model 5: Model 4 + voriconazole on CL | 10585.86 | -330.74 | 10611.90 | -328.7 |
| Model 6: Model 5 + amiodarone on CL | 10541.02 | -44.85 | 10569.00 | -42.9 |
| Model 7: Model 6 + CYP3A4 genotype on CL | 10539.65 | -1.36 | 10569.70 | 0.70 |
| Model 8: Model 6 + fluconazole on CL | 10537.95 | -3.06 | 10567.60 | -1.40 |
| Model 9: Model 6 + total body weight on CL | 10532.47 | -8.55 | 10562.50 | -6.50 |
| Model 10: Model 9 + PGD on CL | 10528.97 | -3.50 | 10562.60 | 0.10 |
| Model 11: Model 9 + total body weight on Vd | 10515.47 | -17.00 | 10547.50 | -15.00 |
| **Multivariable models: backward selection** |  |  |  |  |
| Model 12: Model 11 – total body weight on Vd | 10523.93 | 8.46 | 10553.90 | 6.40 |
| Model 13: Model 11 – amiodaone on CL | 10561.03 | 45.55 | 10591.00 | 43.50 |
| Model 14: Model 11 – voriconazole on CL | 10853.49 | 338.01 | 10883.50 | 336.00 |
| Model 15: Model 11 – CYP3A5 genotype on CL | 10552.36 | 36.89 | 10582.40 | 34.90 |
| Model 16: Model 11 – transplant type on CL | 10590.46 | 74.99 | 10618.50 | 71.00 |
| Model 17: Model 11 – hematocrit | 10645.67 | 130.20 | 10675.70 | 128.20 |
| Model 18: Model 11 – postoperative day on CL | 11153.97 | 638.49 | 11184.00 | 636.50 |

#### Final model evaluation and validation

Diagnostic plots of the final model are shown in Figure 4 (derivation cohort) and Figure 5 (validation cohort). In both cohorts, the pattern of prediction error is more symmetric around the line of identity compared to the base model, but predicted concentrations again underestimated observed values to some extent in the early postoperative time period. In the validation cohort, the final model showed a mean PE of 36.4% (95%CI 30.8%−41.9%) and a median PE of 7.2% (IQR −29.3%−70.53%). The percentage of population predicted concentrations that were within 2 ng/mL or 4 ng/mL of observed was 34.7% and 59.1%, respectively.

**Figure 4.**
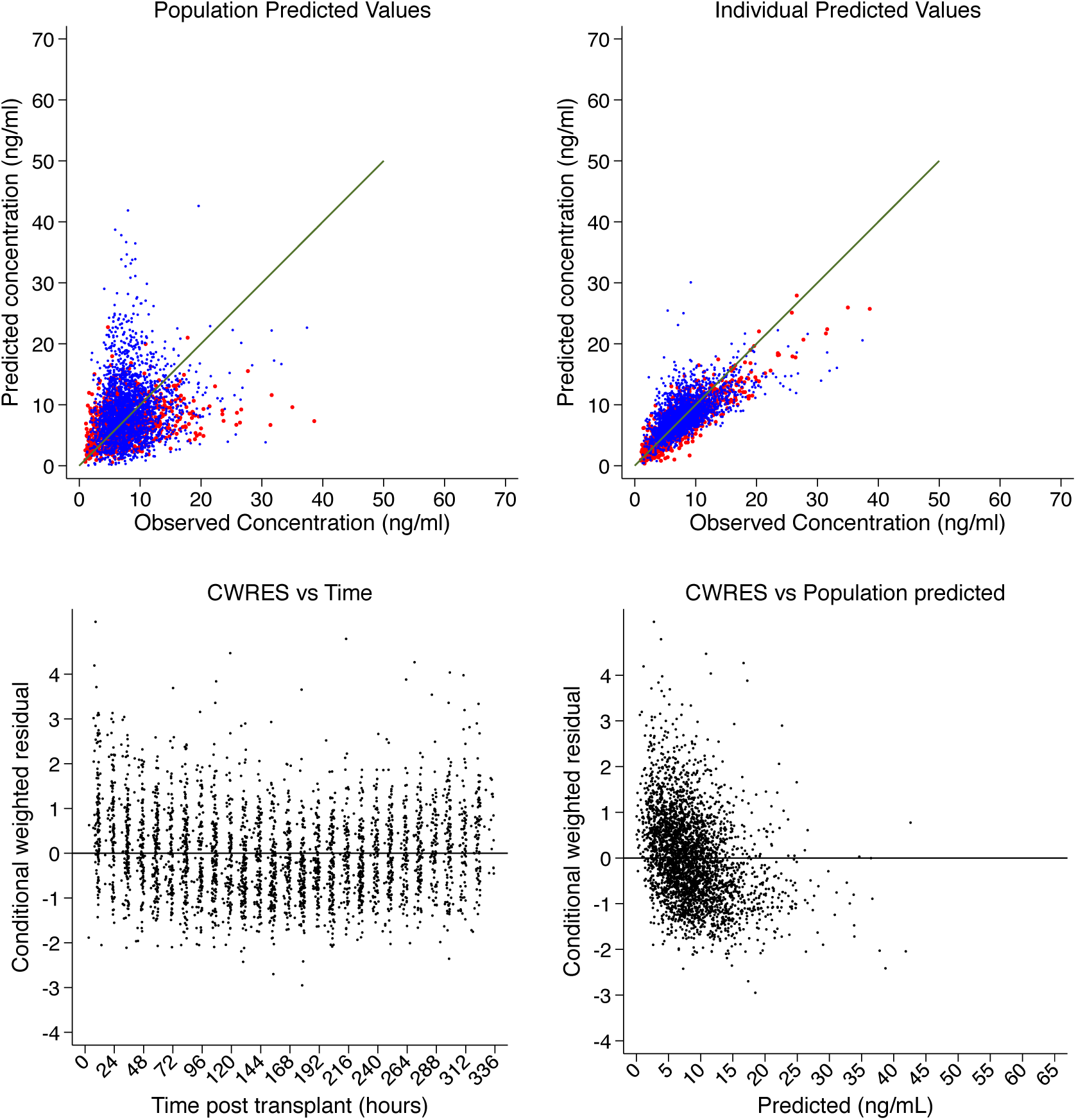
Box plot of population predicted clearance stratified by postoperative day.

**Figure 5.**
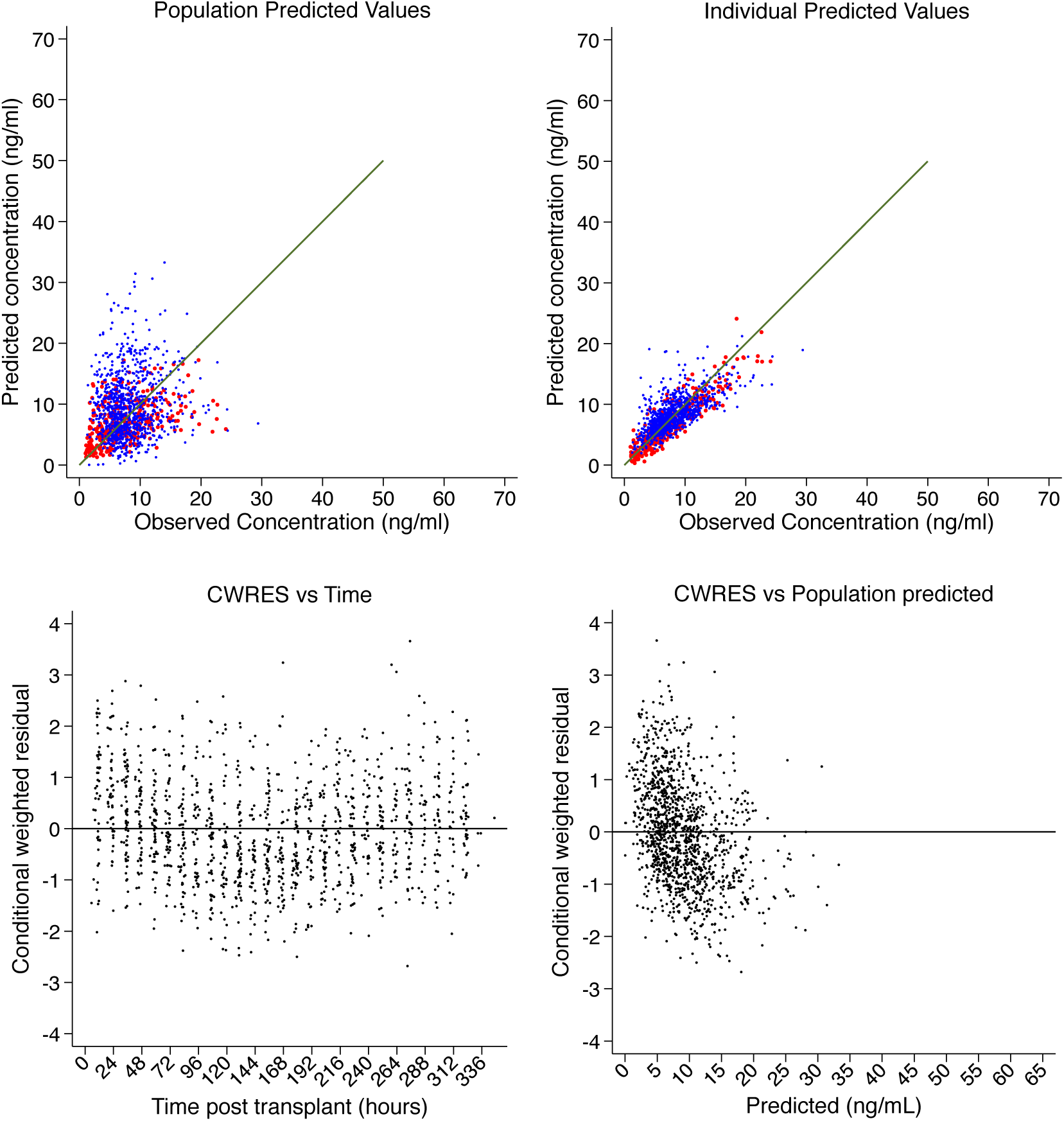
Diagnostic plots of the final model in the validation cohort. Observed vs. predicted plots stratified by postoperative day (red days 0-3, blue days 4-14). CWRES-conditional weighted residual error.

## Discussion

In this study, we leveraged tacrolimus trough concentration data obtained from routine clinical practice to derive and validate a population pharmacokinetic model tailored to the early postoperative time period after lung transplantation. In addition to CYP3A5 genotype, our final model included key clinical factors relevant to lung transplantation such as transplant type and commonly encountered drug-drug interactions. Nevertheless, we observed substantial prediction error when our model was applied to the validation cohort. More comprehensive assessment of clinical covariates in studies employing rich interdose sampling are needed to better explain tacrolimus exposure variability and create a clinically usable population PK model in the early post-lung transplant time period.

Our findings add substantially to the one prior study evaluating tacrolimus PK in postoperative lung transplant patients. Sikma *et al.* developed a tacrolimus population pharmacokinetic model in 20 lung and 10 heart recipients, but noted high interoccurrence variability and concluded that a practice change from oral to intravenous tacrolimus administration should be considered (3). While that study had rich interdose tacrolimus measurements, no clinical covariates were included in the final model and key covariates such as daily hematocrit levels and CYP genotyping were not measured. Although not definitive, our study was larger (n=384) and included multiple clinical covariates including CYP genotype and daily hematocrit, which explained a non-trivial amount of tacrolimus level variability. Despite our model being limited to use of clinically available trough concentrations, validation testing showed that nearly 60% of predicted levels were within 4 ng/ml of observed. While this degree of precision may not support clinical use, our findings clearly raise the possibility that a better-powered study pairing serial interdose tacrolimus measurements with detailed daily clinical data could produce a model with clinical utility. Nevertheless, this study demonstrates that population covariates might be used to improve individual dosing, though more rich sampling is needed to define a robust model. Further, it is highly plausible that a model-based data-driven approach incorporating relevant covariates specific to lung transplantation would improve on the current “trial and error” approach to tacrolimus dosing used perioperatively for most lung transplant recipients.

A key observation from this study is the dynamic nature of tacrolimus clearance during the early period after lung transplantation. Postoperative day was the strongest predictor of clearance, with predicted clearance increasing more than threefold over the study period. We hypothesize that postoperative day is a surrogate for changing critical illness physiology over time as patients recover from the surgical insult (11). We hypothesized that transplant type and PGD would explain some of this underlying physiology. Transplant type is a marker of the severity of the surgical insult, with bilateral transplants requiring longer duration of cardiopulmonary bypass, a median sternotomy, and potentially increased risk for ischemia reperfusion injury (27). PGD is a severe lung injury syndrome that occurs within the initial three days after lung transplantation, is characterized by circulating markers of physiologic dysregulation, and is a strong risk factor for early mortality (28). Notably, these covariates had associations with tacrolimus clearance that were different on postoperative days 0-3 versus later days (although PGD was not significant in multivariable analysis). The time-varying association between transplant type and clearance is consistent with an early impact of the surgical insult that wanes over time as patients recover. This highlights the importance of exporing time varying coefficients for such variables in this population, as the relative lack of association with clearance in the later time period may diminish these variables’ predictive utility. Future analyses that include additional covariates related to critical illness, such as mechanical ventilation parameters, blood pressure, fluid and blood product resuscitation, and intensity of vasopressor support may improve characterization of the effects of critical illness on tacrolimus pharmacokinetics.

In agreement with previous studies (10), we identified *CYP3A5* genotype as a strong predictor of tacrolimus clearance. One in five patients had a *CYP3A5* intermediate or high expresser genotype in our cohort. CYP3A5 expression is more common in African Americans (10), thus the impact of this covariate could be larger in other study cohorts. Although CYP3A4 poor metabolizer genotype has been associated with reduced tacrolimus clearance in previous studies (18,19), it did not demonstrate a significant impact in multivariable PK analysis, which may be due in part to the low number of patients with this genotype in our cohort.

We additionally identified voriconazole and amiodarone as concomitantly administered drugs associated with significantly reduced tacrolimus metabolism. These drugs are known strong inhibitors of CYP3A4, the predominant CYP enzyme responsible for tacrolimus metabolism in those not expressing CYP3A5. Both drugs are key therapies in the lung transplant population, being used more commonly in this group compared to other transplant populations. Voriconazole is commonly used for prophylaxis of invasive aspergillosis (29), and amiodarone is used for the prevention and treatment of postoperative atrial fibrillation, which is common after lung transplantation (30). These covariates were modeled as time varying covariates, lagged by 24 hours. The 24-hour lag specification was based on the timing of routine concentration monitoring in our cohort – daily trough levels are obtained prior to the morning dose (typically given at 6 am). Thus, when an inhibitor drug is initiated, the effects generally will not be observed until the following day’s tacrolimus concentration.

Our study has limitations. The primary limitation is that the timing of tacrolimus concentrations were limited to trough sampling. This limited our ability to explore two-compartment models and to model the absorption process. The latter limitation is particularly important, as tacrolimus absorption is highly variable, and would likely be substantially deranged in the setting of critical illness (3). Lastly, although we examined our final model in a validation cohort, this cohort consisted of other patients transplanted at the same center. Subsequent efforts to produce a robust population pharmacokinetic model for early post-lung transplant tacrolimus dosing should include external validation by incorporating additional centers.

## Conclusion

Tacrolimus exposure and pharmacokinetics are highly variable after lung transplantation, potentially contributing to adverse outcomes in this vulnerable population. This analysis identified several important covariates predicting early postoperative tacrolimus pharmacokinetics, produced a population pharmacokinetic model with fair predictive characteristics, and suggests that future studies employing intensive sampling and detailing time-varying clinical covariates may advance the goal of a clinically usable model in this population.

## Data Availability

Data will be made available on reasonable request.

## References

1. Annual Data Report. Scientific Registry of Transplant Recipients. Available at: http://srtr.transplant.hrsa.gov/annual_reports/Default.aspx. Accessed June 2023

2. Farrell CL, Miano TA, Griffiths S, Christie JD, Diamond JM, Shashaty MGS. Early post-lung transplant calcineurin inhibitor management varies widely: An international survey. Clin Transplant. 2022 Feb;36(2):e14510

3. Sikma MA, Hunault CC, Van Maarseveen EM, Huitema ADR, Van de Graaf EA, Kirkels JH, Verhaar MC, Grutters JC, Kesecioglu J, De Lange DW. High Variability of Whole-Blood Tacrolimus Pharmacokinetics Early After Thoracic Organ Transplantation. Eur J Drug Metab Pharmacokinet. 2020 Feb;45(1):123–134

4. Miano TA, Flesch JD, Feng R, Forker CM, Brown M, Oyster M, Kalman L, Rushefski M, Cantu E 3rd, Porteus M, Yang W, Localio AR, Diamond JM, Christie JD, Shashaty MGS. Early Tacrolimus Concentrations After Lung Transplant Are Predicted by Combined Clinical and Genetic Factors and Associated With Acute Kidney Injury. Clin Pharmacol Ther. 2020 Feb;107(2):462–470

5. Naesens, M., Kuypers, D.R. & Sarwal, M. Calcineurin inhibitor nephrotoxicity. Clin. J. Am. Soc. Nephrol. 4, 481–508 (2009)

6. Wehbe, E., Duncan, a.E., Dar, G., Budev, M. & Stephany, B. Recovery from aKI and short- and long-term outcomes after lung transplantation. Clin. J. Am. Soc. Nephrol. 8, 19–25 (2013)

7. Shashaty MGS, Forker CM, Miano TA, Wu Q, Yang W, Oyster ML, Porteous MK, Cantu EE 3rd, Diamond JM, Christie JD. The association of post-lung transplant acute kidney injury with mortality is independent of primary graft dysfunction: A cohort study. Clin Transplant. 2019 Oct;33(10):e13678

8. Chambers, D. C. et al. The Registry of the International Society for Heart and Lung Transplantation: Thirty-fourth adult lung and heart-lung transplantation report-2017; focus theme: Allograft ischemic time. J. Heart Lung Transpl. 36, 1047–1059 (2017)

9. Todd, J. L. et al. Risk factors for acute rejection in the first year after lung transplant. A multicenter study. Am. J. Respir. Crit. Care Med. 202, 576–585 (2020)

10. Birdwell, K.a., et al. Clinical pharmacogenetics implementation consortium (CPIC) guidelines for CyP3a5 genotype and tacrolimus dosing. Clin. Pharmacol. Ther. 98, 19– 24 (2015)

11. Sikma, M.a., et al. Pharmacokinetics and toxicity of tacrolimus early after heart and lung transplantation. Am. J. Transplant. 15, 2301–2313 (2015)

12. Saint-Marcoux F, Knoop C, Debord J, Thiry P, Rousseau A, Estenne M, Marquet P. Pharmacokinetic study of tacrolimus in cystic fibrosis and non-cystic fibrosis lung transplant patients and design of Bayesian estimators using limited sampling strategies. Clin Pharmacokinet. 2005;44(12):1317–28

13. Monchaud C, de Winter BC, Knoop C, Estenne M, Reynaud-Gaubert M, Pison C, Stern M, Kessler R, Guillemain R, Marquet P, Rousseau A. Population pharmacokinetic modelling and design of a Bayesian estimator for therapeutic drug monitoring of tacrolimus in lung transplantation. Clin Pharmacokinet. 2012 Mar 1;51(3):175–86

14. Diamond, J.M. et al. Clinical risk factors for primary graft dysfunction after lung transplantation. Am. J. Respir. Crit. Care Med. 187, 527–534 (2013)

15. Poquette MA, Lensmeyer GL, Doran TC. Effective use of liquid chromatography-mass spectrometry (LC/MS) in the routine clinical laboratory for monitoring sirolimus, tacrolimus, and cyclosporine. Therapeutic drug monitoring. 2005; 27: 144–50

16. Kirubakaran R, Stocker SL, Hennig S, Day RO, Carland JE. Population Pharmacokinetic Models of Tacrolimus in Adult Transplant Recipients: A Systematic Review. Clin Pharmacokinet. 2020 Nov;59(11):1357–1392

17. Applied Biosystems Axiom 2.0 Reagents: https://www.thermofisher.com/order/catalog/product/901758

18. Tang JT, Andrews LM, van Gelder T, Shi YY, van Schaik RH, Wang LL, Hesselink DA. Pharmacogenetic aspects of the use of tacrolimus in renal transplantation: recent developments and ethnic considerations. Expert Opin Drug Metab Toxicol. 2016 May;12(5):555–65.

19. Elens L, Capron A, van Schaik RH, et al. Impact of CYP3A4*22 allele on tacrolimus pharmacokinetics in early period after renal transplantation: toward updated genotype-based dosage guidelines. Ther Drug Monit. 2013;35(5):608–616

20. Kay, E. Kwateng, F. Geraghty, G. Morgan: Uptake of FK506 by lymphocytes and erythrocytes. Transplant. Proc. 1991; 23: 2760–2762

21. Chow FS, Piekoszewski W, Jusko WJ. Effect of hematocrit and albumin concentration on hepatic clearance of tacrolimus (FK506) during rabbit liver perfusion. Drug Metabolism and Disposition. 1997; 25: 610–616

22. Christie JD, Bellamy S, Ware LB, Lederer D, Hadjiliadis D, Lee J, Robinson N, Localio AR, Wille K, Lama V, Palmer S, Orens J, Weinacker A, Crespo M, Demissie E, Kimmel SE, Kawut SM. Construct validity of the definition of primary graft dysfunction after lung transplantation. J Heart Lung Transplant. 2010 Nov;29(11):1231–9

23. Anglicheau D, Flamant M, Schlageter MH et al. Pharmacokinetic interaction between corticosteroids and tacrolimus after renal transplantation. Nephrol Dial Transplant 2003; 18: 2409–14

24. Mould DR, Upton RN. Basic concepts in population modeling, simulation, and model-based drug development-part 2: introduction to pharmacokinetic modeling methods. CPT Pharmacometrics Syst Pharmacol. 2013 Apr 17;2(4):e38

25. Karlsson MO, Savic RM. Diagnosing model diagnostics. Clin Pharmacol Ther 2007; 82: 17–20

26. Sheiner LB, Beal SL. Some suggestions for measuring predictive performance. J Pharmacokinet Biopharm. 1981;9:503–512

27. Puri V, Patterson GA, Meyers BF. Single versus bilateral lung transplantation: do guidelines exist? Thorac Surg Clin. 2015;25(1):47–54. doi: 10.1016/j.thorsurg.2014.09.007. PMID: 25430429; PMCID: PMC4247525.

28. Christie JD, Kotloff RM, Ahya VN, Tino G, Pochettino A, Gaughan C, DeMissie E, Kimmel SE. The effect of primary graft dysfunction on survival after lung transplantation. Am J Respir Crit Care Med. 2005 Jun 1;171(11):1312–6

29. Marinelli T, Davoudi S, Foroutan F, Orchanian-Cheff A, Husain S. Antifungal prophylaxis in adult lung transplant recipients: Uncertainty despite 30 years of experience. A systematic review of the literature and network meta-analysis. Transpl Infect Dis. 2022 Jun;24(3):e13832

30. Nielsen TD, Bahnson T, Davis RD, Palmer SM. Atrial fibrillation after pulmonary transplant. Chest. 2004 Aug;126(2):496–500

